# How Timing of Stay-at-home Orders and Mobility Reductions Impacted First-Wave COVID-19 Deaths in US Counties

**DOI:** 10.1101/2020.11.24.20238055

**Authors:** Michelle Audirac, Mauricio Tec, Lauren Ancel Meyers, Spencer Fox, Cory Zigler

## Abstract

As SARS-CoV-2 transmission continues to evolve, understanding how location-specific variations in non-pharmaceutical interventions and behaviors contributed to disease transmission during the initial epidemic wave will be key for future control strategies. We offer a rigorous statistical analysis of the relative effectiveness of the timing of both official stay-at-home orders and population mobility reductions during the initial stage of the US epidemic. We use a Bayesian hierarchical regression to fit county-level mortality data from the first case on Jan 21 2020 through Apr 20 2020 and quantify associations between the timing of stay-at-home orders and population mobility with epidemic control. We find that among 882 counties with an early local epidemic, a 10-day delay in the enactment of stay-at-home orders would have been associated with 14,700 additional deaths by Apr 20 (95% credible interval: 9,100, 21,500), whereas shifting orders 10 days earlier would have been associated with nearly 15,700 fewer lives lost (95% credible interval: 11,350, 18,950). Analogous estimates are available for reductions in mobility—which typically occurred before stay-at-home orders—and are also stratified by county urbanicity, showing significant heterogeneity. Results underscore the importance of timely policy and behavioral action for early-stage epidemic control.

---

SARS-CoV-2, the causative virus of COVID-19, continues to threaten the world with nearly 9.2M reported cases and 348,000 deaths as of Jan 1st, 2021 in the US^1^. It is clear that both non-pharmaceutical interventions (NPIs) and behavioral changes have impacted transmission of the virus^2–7^. However, there are multiple population-specific factors that influence epidemic trajectories and how they change including demographic characteristics and, importantly, the timing of such NPIs and behavioral responses^8,9^. Decoupling the impacts of these factors and the relative timing of actions or behavior changes presents important challenges^5,10^. A key problem particularly in the United States, is that policy changes and mobility shifts happened rapidly in late March and April 2020 in response to growing COVID-19 epidemic and happened simultaneously across the country, so it has thus far been difficult to disentangle the relative impact of earlier vs. later action and behavioral responses from NPIs.

In this work, we quantify the impact of the timing of stay-at-home orders and mobility reduction relative to local epidemic conditions. Specifically, we developed a spatial Bayesian statistical model of county-level COVID-19 mortality between January 21, 2020 and Apr 20, 2020 across 882 counties, most of which had either: a) an official stay-at-home order in place or b) a reduction in population mobility of at least 50% relative to baseline during this time period. We model how county-specific trajectories of COVID-19 deaths changed as a function of timing of control efforts and county-level features and use the model to predict county-specific death trajectories under alternative timing of stay-at-home orders or mobility reductions.

## METHODS

### County-level COVID death and demographic data

Our analysis relies on multiple county-level data sources. First is the dataset compiled by Kileen et al.^11^ reported by the Center for Systems Science and Engineering (CSSE) at Johns Hopkins University, which counts COVID-19 related deaths as well as the dates in which different NPIs took place in each county. Figure S2 in the Web Appendix shows a comparison of the dates at which different NPIs were enacted. For our statistical model, we focus specifically on stay-at-home orders for two reasons: first, they exhibit wide variation in timing among counties; second, counties typically had sufficiently long periods of COVID-19 deaths pre-intervention, a condition useful for statistical estimation of pre-vs. post-intervention trajectories. For instance, 56% of the counties had at least 10 pre-intervention daily observations before the stay-at-home orders (considering the delay from infection to death). In contrast, for the five policies contained in the data but not included in the study, less than 25% of the counties show pre-intervention periods of at least 10 days.

A second source of data pertains to county-level covariates, which are included in the analysis with two objectives in mind: 1) to reflect the differences in disease dynamics associated with different types of counties and 2) control for possible differences between counties with different timing of stay-at-home orders or mobility reductions. As a summary measure of county characteristics known to relate to residents’ contact rates and the reproduction number (population density, modes of travel, distance to major airports) and other factors expected to vary across the spectrum of rural and urban areas, we use the National Center for Health Statistics (NCHS) Urban-Rural Classification Scheme for Counties^12^, which classifies each US county to be in one of the following six categories: 1) large central metro; 2) large fringe metro; 3) medium metro; 4) small metro; 5) micropolitan; 6) non-core. In addition, we extract from the US Census American Community Survey other county-level covariates that have been reported to have a strong relationship with COVID-19 death rate and are not captured by the NCHS county classification. These covariates are further described in the Methods section.

### Human mobility data

Focusing on a single well-defined intervention, here, the stay-home orders, is not able to parse the possible contributing effects of other concurrent or overlapping early-epidemic control efforts. For this reason, we also investigate the timing of a mobility-related behavior change to capture policy-induced or voluntary behavioral responses to the epidemic. Thus, for our third data source we use human mobility data from SafeGraph, a company that provides anonymized population mobility information representing 45 million smartphone devices. SafeGraph data aggregates visit counts to numerous points of interest (POIs) classified into categories. As a proxy measure for overall mobility behavior in each county, we extract data (accessed from Safegraph in May, 2020) on the number of visits per day to POIs; in particular to schools, colleges, restaurants, bars, parks and museums, and obtained time series of daily total visits for each county. The average total visits per day between Jan 15, 2020 and Feb 15, 2020 is used to establish county-baseline levels of mobility. We define the date of the mobility-based intervention to be the date on which the right-aligned ten-day moving average of total visits to POIs decreased 50% relative to baseline. All counties in the data set reached at least a 40% mobility reduction, with a few counties reducing by more than 75%. After mobility was substantially decreased, it held fairly constant during the analysis time frame. Therefore, the date at which the 50% cut-off was reached is chosen as a practical balance to represent the start of a period of low levels of visits to POIs.

### Analysis data set

County-specific daily counts of COVID-19 deaths were extracted from Jan 21, 2020 through April 20, 2020 which we take to be a loose definition of the end of the first US epidemic wave, as mobility started to rise again after this date. During that period, 1243 counties (out of a total of 3221 US counties) had reached a threshold of 3 deaths per 10 million residents, signaling arrival of a local epidemic. We restrict attention to those counties that have at least seven days recorded after reaching their threshold and excluded 222 counties due to lack of sufficient Safegraph mobility data to characterize baseline mobility. A map in Figure 1 depicts the 882 counties of interest that comprise the analysis dataset in which 93.4% instituted stay-at-home orders, and 99% exhibited at least 50% mobility reductions. Table S2 in the Web Supplement summarizes the number of counties and population covered in each NCHS classification category, indicating that this fraction of US counties in the dataset covers approximately 234 million, roughly 71.5% of the total US population.

**Figure 1:**
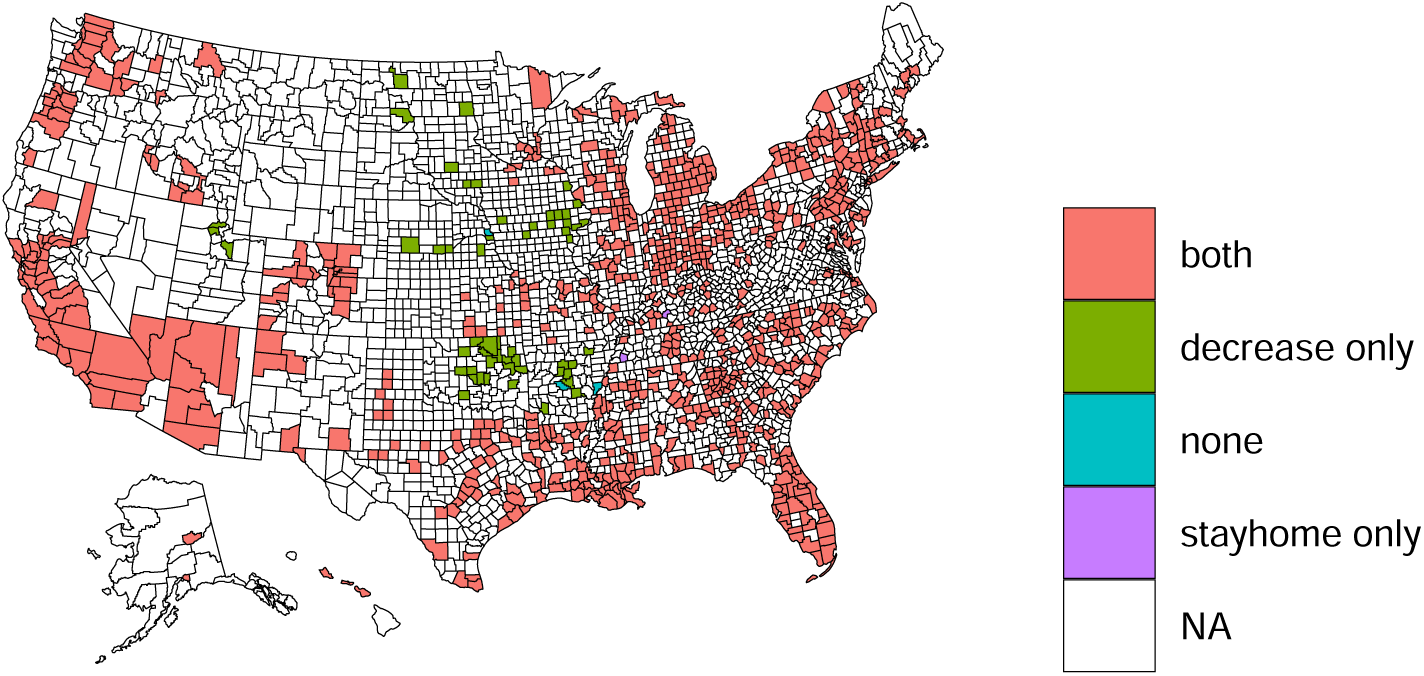
Geographic representation of 882 counties comprising the analysis dataset: 822 had both interventions of interest, 58 only mobility decrease, 5 only stay-home orders, and 3 none of the two interventions. NA stands for counties that did not reach the death threshold, or did not adhere to the minimum required number of records of information in the analysis time period. Additional details on counties coverage are found in Table S2.

### Statistical methods

We propose a spatial Bayesian hierarchical model to capture the impact of the timing of an intervention (stayhome or mobility decrease) as measured by a change in trend in death trajectories. The analysis consists of two distinct variations: 1) *Stay-at-home model* : Here, the intervention and relative timing are defined according to the date at which a county instituted a stay-at-home order, 2) *Mobility model* : Here the “intervention” is not actually a discrete policy intervention, but the date at which a county reached a 50% reduction from baseline mobility patterns. Throughout, we continue to refer to the mobility reduction as an “intervention” for simplicity.

To model death trajectories, we use the time series of each county’s 7-day centered rolling average of daily deaths, and specify a log-linear model for the expected number of deaths in each county using a negative binomial distribution and a two-stage quadratic function of the “epidemic time”, defined as the number of days elapsed since reaching a deaths threshold of 3 per 10 million residents. The two-stage time function is modeled to change after a pre-specified delay following introduction of an intervention, resulting in different pre- and post-intervention trajectories. A schematic representation of the components of the proposed model appears in Figure 2. Note that this work focuses on estimating the difference between post-intervention trends under different intervention timing, and not on a comparison with the implied extension of the pre-intervention trend through the post-intervention period (which would correspond to no intervention at all).

**Figure 2:**
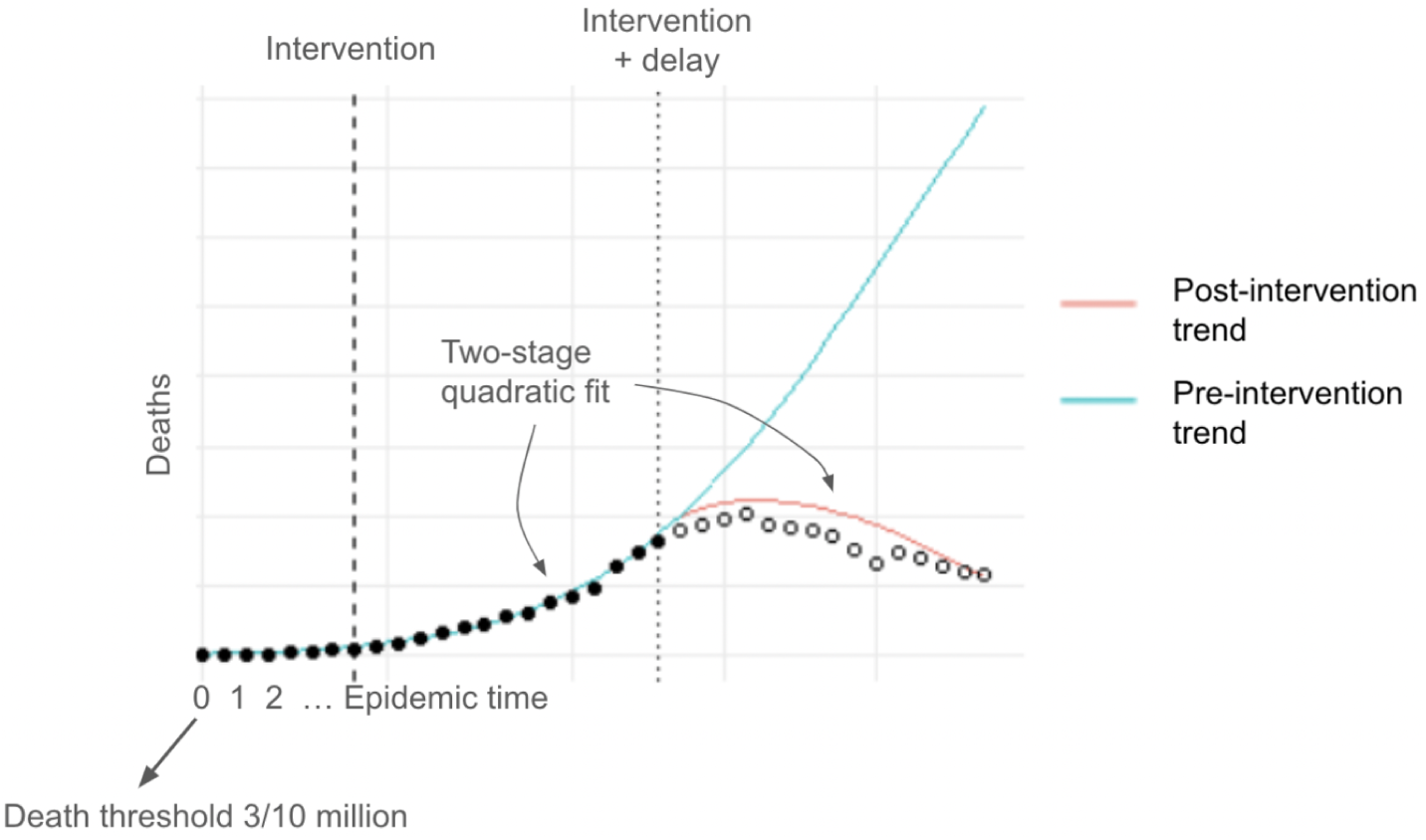
The model specification includes two types of interaction variables. First, are interactions between county demographics and pre-intervention quadratic time terms as described in Web Supplement B. These induce flexibility in modeling pre-intervention disease dynamics in different types of counties. Second is the intervention timing and the post-intervention time terms. Analogous interactions between county features have the role of allowing the change of the post-intervention death trajectory to vary according to county features. These latter post-intervention interactions modify the effect of intervention timing, allowing the degree of “bend” in a death trajectory associated with different timings to vary according to county features.

The ability of quadratic functions to naturally adapt to epidemic curves that start growing exponentially and eventually flatten was particularly useful to the Centers of Disease and Control Prevention during the first-wave of the Covid-19 pandemic when epidemiological parameters remained elusive^13^. The choice of a log-quadratic specification is further explored in Woody et al.^14^. Our approach is similar to theirs in that it modulates the coefficients of the log-quadratic trends, and in that it uses a negative binomial likelihood and county-level random effects. The model utilizes a lag of 14 days after introduction of an intervention before a change in death trajectories, a delay consistent with the first quartile of the distribution of time between infection and death^15–17^, and informed by an analysis in Web Supplement B.6 that estimates this lag from available data.

To parsimoniously capture variation across counties, a county’s pre-invervention trajectory is modeled to interact with the county’s NCHS county classification as well as county-specific demographics. These are meant solely to induce flexibility in modeling pre-intervention disease dynamics in different types of counties, and these are all features with sufficient reason to suspect that they relate to disease transmission. The percent of black residents and percent of hispanic residents are included to account for the apparent disparities between infection and comorbidity and death rates among the population. To account for the age-related risk of death, we include the percentage of residents that are 65 years or older. Particular behaviors specific to students (whose main activity quickly became completely remote amid the early epidemic stages), are accounted with the percentage of residents attending college.

In addition to demographic covariates, three different types of random effects are included to model latent heterogeneity in the polynomial terms of the pre-intervention trajectories: two sets of county-level random effects model a) spatially-varying heterogeneity based on the Besag-York-Molli é formulation^18^ to capture correlation between neighboring counties and b) additional heterogeneity with unstructured random effects. A third set of state-level random effects model correlation among counties within the same state that adhere to state-level policies.

For the post-intervention trajectories, the same demographic factors are used as well as their interaction with time polynomials, augmented with an explicit term for the timing of the intervention. These covariates have the analogous role of allowing the change of the post-intervention death trajectory to vary or “bend” according to county features, modeling different deaths trajectories under different intervention timings.

Refer to Web Appendix B for a detailed mathematical formulation of the model and specification of the parameters’ priors; inferences are based on posterior simulations from the models. We fit all models using the R language (4.1) with the package rstan (2.19.3)^19^. Processed data, model code, and the resulting data sets are available with no restrictions at https://github.com/audiracmichelle/covid_timing.

## RESULTS

### Epidemic timing

Among the 882 counties in the present analysis, death thresholds of 3 per 10 million residents were reached at different calendar times with differences across levels of NCHS county classification shown in Figure 3. According to the average calendar date, the epidemic arrived earliest in large central metro areas. In terms of “epidemic timing,” the median and IQR of the number of days between a county deaths threshold and the issuing of the stay-at-home order was -3 (−9, 2), and -9 (−15, -4) for 50% mobility reductions. As shown in Figure 4, the epidemic timing of stay-at-home orders exhibited the opposite temporal ordering of epidemic arrival with stay-at-home orders occurring earliest in epidemic time in non-core and micropolitan counties. The relative epidemic timing at which counties across different NCHS categories reached this mobility reduction showed the same ordering as the dates of stay-at-home orders. Note that mobility reductions occurred earlier, with mobility reductions tending to precede stay-at-home orders by a median of -5 (−10, -2) days across all NCHS categories.

**Figure 3:**
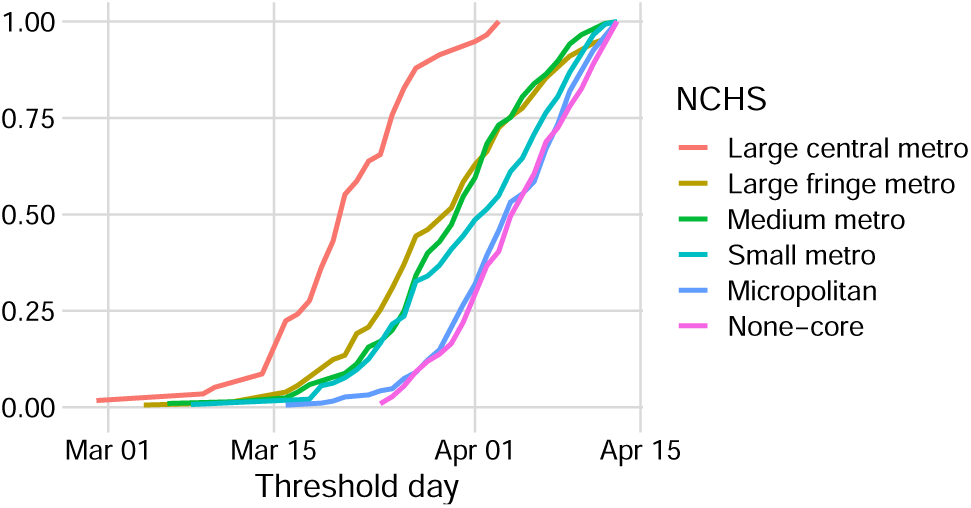
Cumulative percentage of counties of interest (*y*-axis) in each category that had reached the threshold of 3 deaths per 10 million residents by the corresponding calendar date. The epidemic arrived earliest in large central metro areas (average date March 21), followed by large fringe, medium, and small metro areas (average dates March 29, March 30, and Apr 1st, respectively), with the latest epidemic arrival in micropolitan and non-core counties (both with average date April 4).

**Figure 4:**
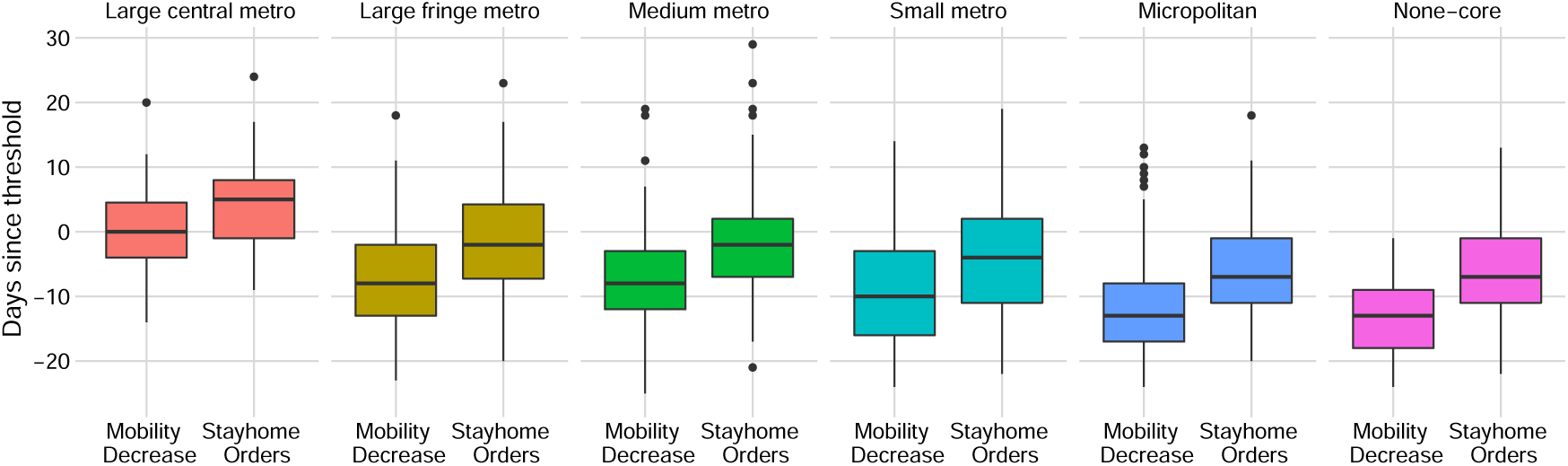
Epidemic timing of mobility reduction and stay-at-home orders across NCHS groups. Days since threshold (*y*-axis) of 0 represents the day that the threshold of 3 deaths per 10M residents was reached, negative numbers represent days before the threshold. The specific values for the median, 25%tile and 75% tile are found in Table S2 in Web Supplement. The distribution of mobility reductions timing in general occurred earlier than stay-home orders, and almost always before a county even reached the 3/10 million death thresholds in all NCHS categories except large central metro counties.

### Modeling results

County-specific death trajectories were fitted using the model described in the Methods section. Several diagnostics were performed to assess the adequacy and fit of the model, including MCMC convergence diagnostics, posterior-predictive checks, and prediction an a hold-out sample to verify that inferences were not dominated by a small number of large counties. Full details appear in Web Supplement C, which yield confidence in the parsimonious yet flexible formulation of the proposed model. Web supplement C.3 displays the lack of significant residual spatial autocorrelation. Web Supplement C.6 illustrates the posterior fit for six counties with heterogeneous interventions timing.

### Impact of intervention timing

To evaluate the impact of intervention timing we offer posterior-predictive simulations for each county under hypothetical scenarios where the intervention occurred 10 days earlier or later than observed, tabulating estimated deaths through April 20. Figure 5 shows the results of using the fitted statistical model to compute the *per capita* median trajectory of daily deaths for each NCHS category under the observed intervention scenario and hypothetical intervention timings. The top panel evidences that enacting stay-at-home orders 10 days earlier would have strongly mitigated the trajectory of daily deaths, particularly for the more urban counties, although the impact is noticeable in all NCHS categories. The bottom panel depicts analogous fitted and counterfactual trajectories for the Mobility decrease model. While the general shape of trajectories is similar to those of the stay-at-home interventions, some differences are apparent. For small metro and micropolitan counties, there is more pronounced evidence that earlier mobility intervention impacted the daily death trajectories. In contrast, the evidence of impact for non-core counties is less pronounced for mobility decrease. A plausible explanation is that the these counties tend to have noisy mobility data since there are less POIs and mobile devices registered by Safegraph.

**Figure 5:**
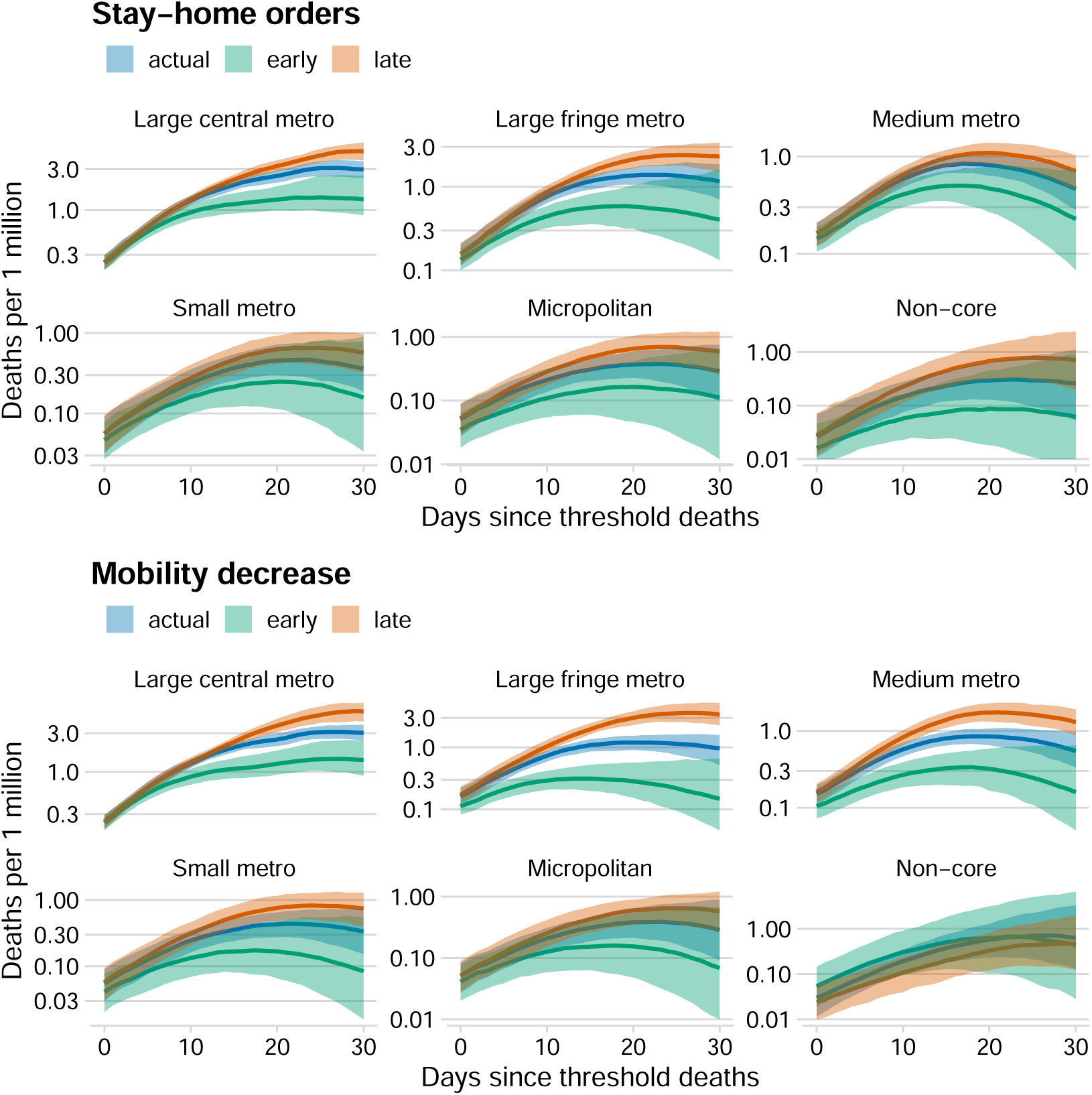
Estimated death trajectories for observed timing, 10-day earlier timing, and 10-day delayed timing of stay-at-home orders and mobility reductions. Solid lines are posterior median estimates of deaths per 1 million residents at *t* days after epidemic arrival. Shaded areas are 95% posterior credible intervals.

Table 1 provides estimates of cumulative deaths differences under earlier and later introduction of stay-at-home orders, the same are cast per 100,000 residents. Overall, the model predicts that implementing stay-at-home orders 10 days earlier would have led to 15,700 (95% credible interval: 11,350 18,950) fewer deaths through April 20. Effects are heterogeneous across county classifications with effects concentrated in the large central, large fringe, and medium metro counties. The evidence that delayed action would have led to extra deaths is slightly weaker; adopting stay-at-home orders 10 days later would have led to an additional 14,700 (95%CI: 9,100 21,500) deaths. Table 1 has analogous results for shifting mobility reductions 10 days earlier in these counties which would have averted an estimated 15,550 (95%CI: 10,450 19,400) deaths, while delayed mobility reductions by 10 days would have led to an additional 21,100 (95%CI: 14,500 29,200) deaths over what was observed. In total, the results from the *Mobility model* relative to the *Stay-at-home model* match expectations, since stay-at-home orders typically happened after a significant decrease in mobility had already taken place (*c*.*f*. Figure 4), with mobility drops persisting beyond the first date reaching a 50% reduction from baseline.

**Table 1:**
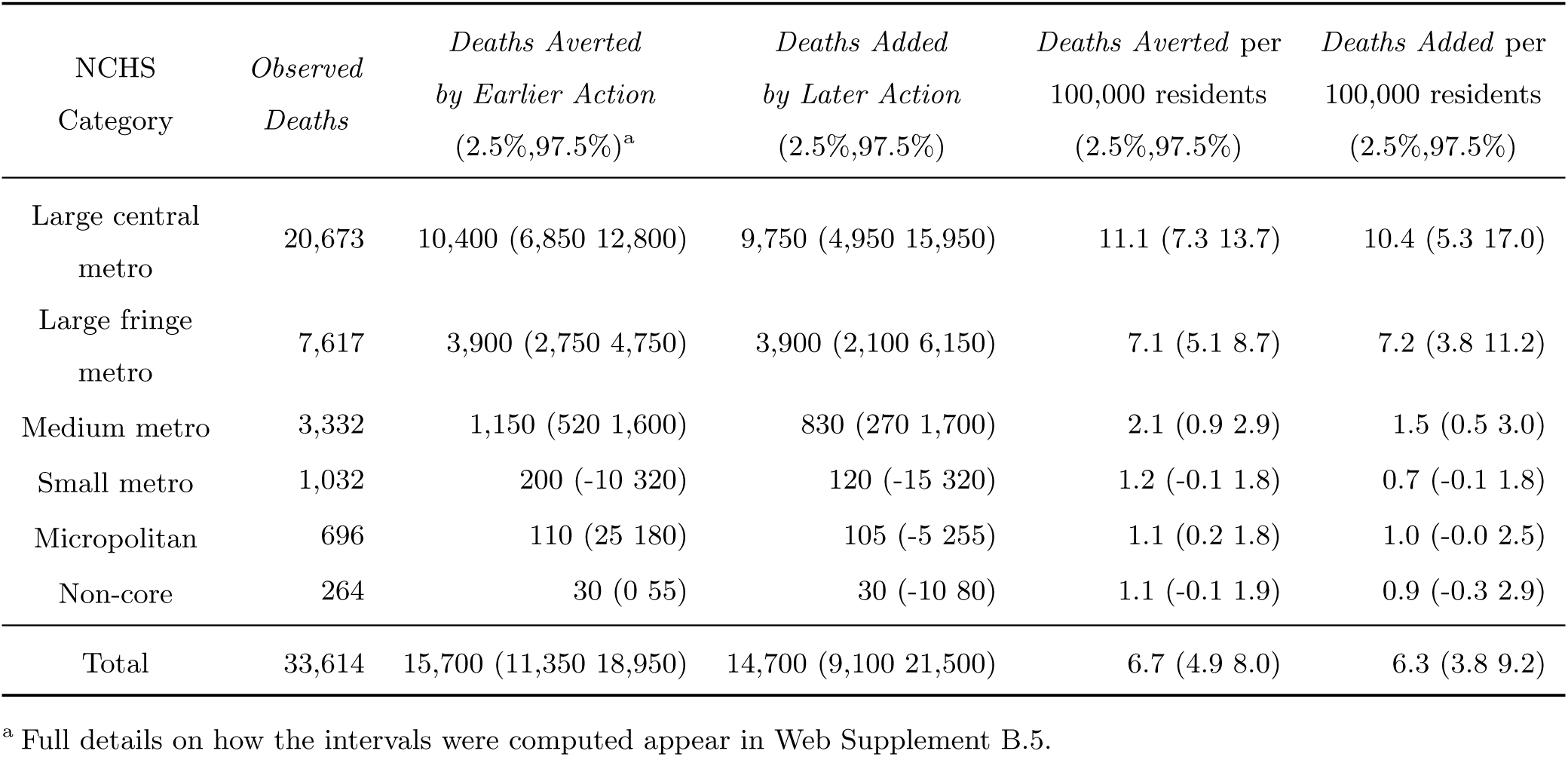
Bayesian posterior estimates using the *Stay-at-home model* for medians and 95% credible intervals of cumulative differences in COVID-19 deaths by 10-day earlier/later action for the 882 counties in the data set from the first death through April 20, 2020.

| NCHS Category | Observed Deaths | Deaths Averted by Earlier Action (2.5%,97.5%)<sup>a</sup> | Deaths Added by Later Action (2.5%,97.5%) | Deaths Averted per 100,000 residents (2.5%,97.5%) | Deaths Added per 100,000 residents (2.5%,97.5%) |
| --- | --- | --- | --- | --- | --- |
| Large central metro | 20,673 | 10,400 (6,850 12,800) | 9,750 (4,950 15,950) | 11.1 (7.3 13.7) | 10.4 (5.3 17.0) |
| Large fringe metro | 7,617 | 3,900 (2,750 4,750) | 3,900 (2,100 6,150) | 7.1 (5.1 8.7) | 7.2 (3.8 11.2) |
| Medium metro | 3,332 | 1,150 (520 1,600) | 830 (270 1,700) | 2.1 (0.9 2.9) | 1.5 (0.5 3.0) |
| Small metro | 1,032 | 200 (-10 320) | 120 (-15 320) | 1.2 (-0.1 1.8) | 0.7 (-0.1 1.8) |
| Micropolitan | 696 | 110 (25 180) | 105 (-5 255) | 1.1 (0.2 1.8) | 1.0 (-0.0 2.5) |
| Non-core | 264 | 30 (0 55) | 30 (-10 80) | 1.1 (-0.1 1.9) | 0.9 (-0.3 2.9) |
| Total | 33,614 | 15,700 (11,350 18,950) | 14,700 (9,100 21,500) | 6.7 (4.9 8.0) | 6.3 (3.8 9.2) |

<sup>a</sup> Full details on how the intervals were computed appear in Web Supplement B.5.

**Table 2:** Bayesian posterior estimates using the *Mobility model* for medians and 95% credible intervals of cumulative differences in COVID-19 deaths by 10-day earlier/later action for the 882 counties in the data set from the first death through April 20, 2020.

| NCHS<br>Category | *Observed*<br>*Deaths* | *Deaths Averted*<br>*by Earlier Action*<br>(2.5%,97.5%)<sup>a</sup> | *Deaths Added*<br>*by Later Action*<br>(2.5%,97.5%) | *Deaths Averted per*<br>*100,000 residents*<br>(2.5%,97.5%) | *Deaths Added per*<br>*100,000 residents*<br>(2.5%,97.5%) |
| --- | --- | --- | --- | --- | --- |
| Large central<br>metro | 20,673 | 9,000 (4,200 12,300) | 9,300 (4,700 15,100) | 9.6 (4.5 13.1) | 9.9 (5.0 16.1) |
| Large fringe<br>metro | 7,617 | 4,800 (3,850 5,500) | 8,500 (5,200 12,350) | 8.7 (7.0 10.0) | 15.5 (9.5 22.6) |
| Medium metro | 3,332 | 1,600 (990 2,000) | 2,650 (1,450 4,050) | 2.9 (1.8 3.6) | 4.7 (2.6 7.3) |
| Small metro | 1,032 | 235 (25 390) | 260 (45 605) | 1.3 (0.1 2.2) | 1.5 (0.3 3.5) |
| Micropolitan | 696 | 80 (-45 160) | 70 (-35 240) | 0.8 (-0.4 1.6) | 0.7 (-0.3 2.4) |
| Non-core | 264 | -40 (-230 20) | -20 (-50 20) | -1.5 (-8.1 0.7) | -0.7 (-1.7 0.7) |
| Total | 33,614 | 15,550 (10,450 19,400) | 21,100 (14,500 29,200) | 6.6 (4.5 8.3) | 9.0 (6.2 12.4) |

<sup>a</sup> Full details on how the intervals were computed appear in Web Supplement B.5.

## DISCUSSION

We have offered a rigorous evaluation of the association between the change in death trajectories and the timing of both stay-at-home orders and reduced population mobility, as measured by number of visits to various POIs. The statistical approach presented here expresses county-specific curves of daily COVID-19 deaths in terms of pre- and a post intervention trajectories, dictated by both observed and latent county-level features and the epidemic timing of the interventions. An important feature of the analysis is that its focus on estimating the impact of early vs. late action, and not on the related impact of implementing vs. not implementing policies or behavior changes.

The descriptive analysis of the timing of interventions clarified both the heterogeneity in intervention timing relative to local epidemic conditions and that mobility reductions often occurred before stay-at-home orders, although not always. By-and-large, in more urban counties the epidemic tended to arrive earlier in calendar time, with policy and mobility interventions in these areas tending to be later in epidemic time relative to less urban counties. Also, the total visits to POIs during the study’s time period had often reached close to its minimum levels at the time an order was instituted. The suite of statistical models fit indicate that in more urban counties the timing of mobility reductions was more important for dictating changes in the daily deaths trajectories than the timing of official stay-at-home orders, while the uncertainty of the results for rural counties are rendered inconclusive. This is not to say that the stay-at-home orders had no impact, and interpretation of the interplay between these two types of “interventions” is not within the scope or the present work.

The modeled associations and death projections under counterfactual intervention timing can be interpreted as causal effects of the intervention timing under the assumption that the functional form of the model correctly characterizes death trajectories and that the model adjusts for relevant confounders that dictate intervention timing and deaths. Web Supplement A.4 evidences the importance of the included confounders for their ability to explain variation in intervention timing, and also indicates no clear threat of unmeasured spatially-varying factors that dictated intervention timing. However, even with the apparent adequate fit of the model and the specific elements of the approach designed to address major threats to causal validity, the implications of the results should be viewed in light of the potential confounding due to unobserved factors might not be fully resolved.

Also worth noting is that the results herein pertain to the 882 counties included in the analysis which, despite containing 71.5% of the total US population, may not represent epidemic and intervention dynamics in other counties not included in the analysis. Included counties were selected primarily on the basis of having a pronounced epidemic during the first US wave, but 222 counties (114 of which reached the deaths threshold of 3 deaths per 10 million residents) were excluded due to lack of complete or inconsistent mobility data from Safegraph, which occurred due to low coverage of mobile phone data on visits to POIs, difficulty in our processing pipeline for linking POIs to county identifiers, or fewer than 100 days of observed mobility data during the study period. Reasons for counties exhibiting any of these difficulties are not clear, but counties omitted due to lack of mobility data spanned every category of NCHS county classification (with 5% of omitted counties in Large Central Metro areas, and 44% of omitted counties in Micropolitan or Non-core counties) and had no discernible geographic pattern. To the extent that disease dynamics might be different in counties with no pronounced epidemic during the first US wave or in counties with missing Safegraph mobility data, the results herein do not necessarily generalize to those counties.

Our work surmounts some of the challenges of disentangling the intertwined local characteristics and events that unfolded around the time of the first wave of COVID-19 interventions. The statistical evidence generated from a model such as this is designed to complement that obtained from more traditional mechanistic epidemic models. Reproducing the full epidemiological cycle is not the intention of the model; we only attempt to quantify intervention impacts in the time frame surrounding observed stay-at-home and mobility reduction dates and during the early epidemic stages. In fact model fit noticeably deteriorated when fit to longer time periods. The limitations of this analysis notwithstanding, we move a step closer into parsing these events by providing evidence for the timing of official policy interventions and mobility-related behavior changes as important determinants of local daily COVID-19 deaths. These results point towards the need to investigate how official reopening policies and other policies that varied across counties interplay with changes in mobility beyond the time frame considered here and into the later phases of the US COVID-19 epidemic.

## Data Availability

Data on COVID-19 deaths were provided by the New York Times and are publicly available: https://github.com/nytimes/covid-19-data/ Data on social-distancing metrics were provided under a Data Use Agreement by SafeGraph and are available by contacting SafeGraph directly: https://www.safegraph.com/dashboard/covid19-shelter-in-place

https://www.safegraph.com

## WEB APPENDIX

### A Exploratory analysis of timing of policies and mobility reductions

#### A.1 Counties of interest

**Table S2:**
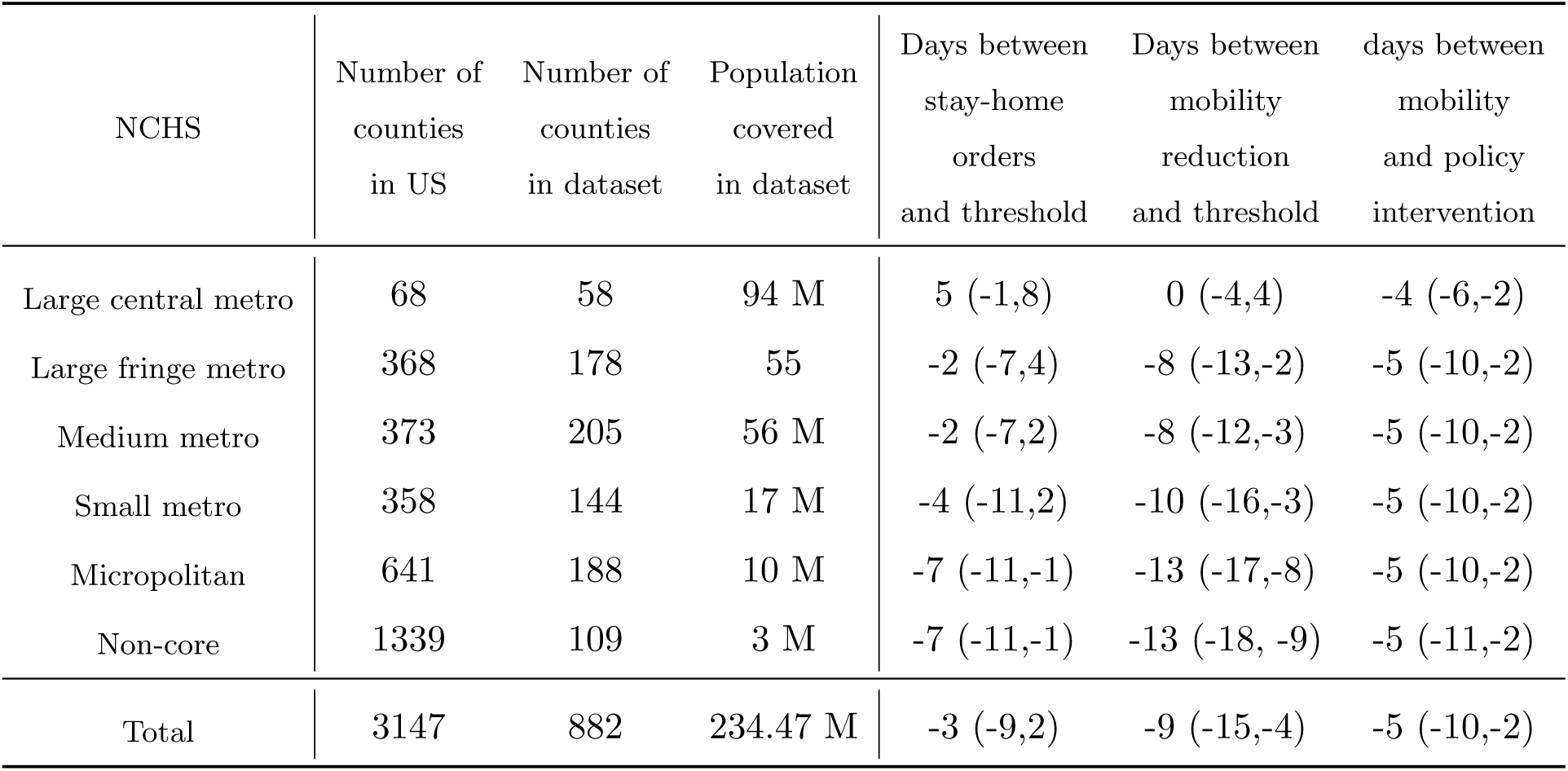
Counties coverage in the analysis grouped by NCHS along with the median number of days between interventions and death threshold. In parenthesis, values for the first and third quantiles are included (25%tile, 75%tile). Stay-at-home orders occurred earliest in epidemic time in non-core counties (median 7 days before deaths threshold), followed by micropolitan counties, small, medium and large fringe metro areas (median days between thresholds and stay-at-home orders of -7, -4, -2, -2, respectively), and large central metro areas having stay-at-home orders latest in epidemic time (median of 5 days after threshold).

| NCHS | Number of<br>counties<br>in US | Number of<br>counties<br>in dataset | Population<br>covered<br>in dataset | Days between<br>stay-home<br>orders<br>and threshold | Days between<br>mobility<br>reduction<br>and threshold | days between<br>mobility<br>and policy<br>intervention |
| --- | --- | --- | --- | --- | --- | --- |
| Large central metro | 68 | 58 | 94 M | 5 (-1,8) | 0 (-4,4) | -4 (-6,-2) |
| Large fringe metro | 368 | 178 | 55 | -2 (-7,4) | -8 (-13,-2) | -5 (-10,-2) |
| Medium metro | 373 | 205 | 56 M | -2 (-7,2) | -8 (-12,-3) | -5 (-10,-2) |
| Small metro | 358 | 144 | 17 M | -4 (-11,2) | -10 (-16,-3) | -5 (-10,-2) |
| Micropolitan | 641 | 188 | 10 M | -7 (-11,-1) | -13 (-17,-8) | -5 (-10,-2) |
| Non-core | 1339 | 109 | 3 M | -7 (-11,-1) | -13 (-18, -9) | -5 (-11,-2) |
| Total | 3147 | 882 | 234.47 M | -3 (-9,2) | -9 (-15,-4) | -5 (-10,-2) |

#### A.2 Policy interventions

Amid the initial spread of COVID-19, an array of non-pharmaceutical interventions including bans on gatherings, closures of schools and restaurants, and stay-at-home orders were implemented. Intervention policies were most-often put in place state wide, but there are cases in which counties adopted such measures before their respective states. Generally speaking, orders to stay at home came last in a quick succession of policies that occurred primarily between March 15 and April 1st. Most bans and closures occurred in the same small window of time and were preceded by stay-at-home orders. Figure S2 shows the number of counties that adopted each of the six policy interventions on a given day.

**Figure S2:**
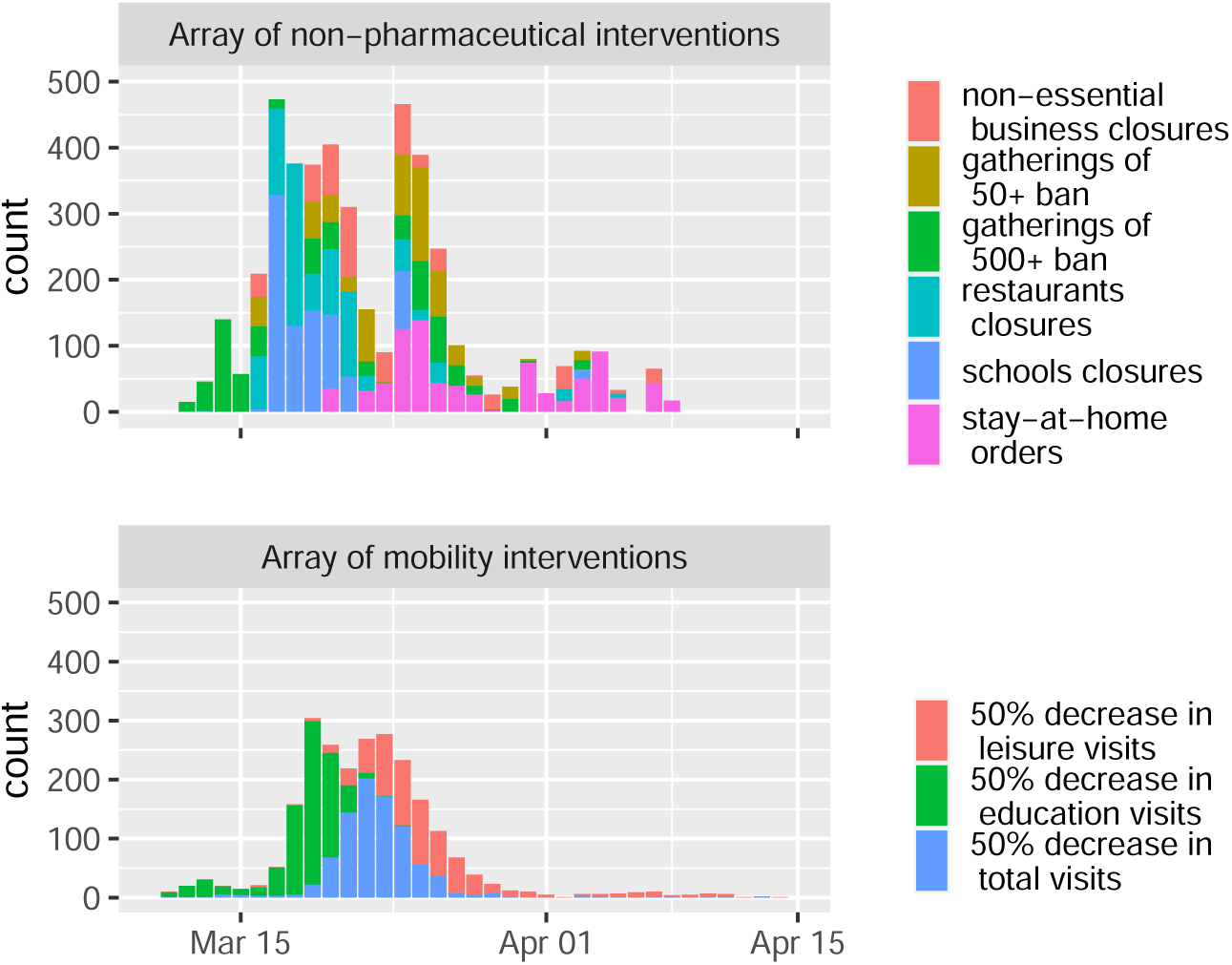
Adoption timeline of an array of both policy and mobility interventions. On the top, counts of dates for six different NPIs, such as stay-home orders, school closures and other more lenient measures, are stacked. On the bottom, counts of days when total visits reached a 50% decrease are included. To illustrate the components that make up total visits, the days “education” and “leisure” visits decreased 50% are also shown; with the education classification covering schools and colleges, and leisure covering restaurants, bars, parks and museums.

#### A.3 Mobility decrease

Most counties exhibited a steady decrease in mobility within a window of ten days, with March 22 being the median date at which counties reached the 50% decrease from their baseline visits to all POIs, corresponding to the definition of a mobility-based intervention. Figure S2 shows the dates when counties achieved a 50% decrease in visits to two sub-categories of POIs: 1) schools and colleges and 2) leisure destinations such as restaurants, bars, parks and museums. Safegraph provides visits data for several other types of POIs, as well as data on the number of minutes devices remain at home or at work, we limited our analysis only to those categories that had a comprehensive coverage across all the range of counties.

**Figure S3:**
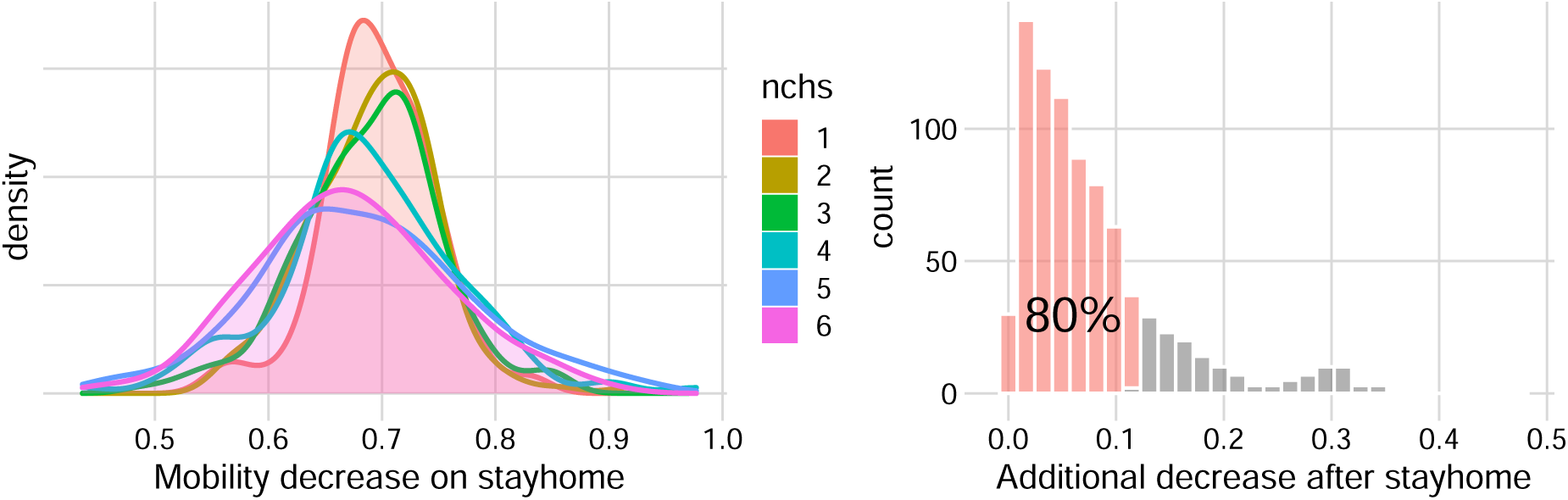
Left: Shows the relative decrease in mobility with respect to the baseline on the day the stay-at-home order was enacted. Right: Shows the additional decrease in mobility since stay-home up to each county’s minimum mobility level. 80% of counties decreased at most 10% after the stay-home orders.

To further illustrate how the timing of mobility interventions compared to that of policy interventions among the 822 counties with both, Figure S3 depicts the decrease in mobility (relative to baseline) that was observed on the day a stay-at-home order took effect. All counties had already shown a marked reduction, with micropolitan and non-core counties showing a reduction of 66% on the day of the policy, and other more urban counties having even more reduced mobility, with an average reduction of 70% mobility when the stay-at-home order took effect.

Along with this, reductions in mobility were close to a level of saturation when stay-home went into effect with total visits to POI not reducing much more. The distribution on the right panel in Figure S3 shows that for 80% of the counties in our dataset, the mobility levels observed the day stay-home stay-home orders were adopted, were at most 10% above their absolute minimum levels.

#### A.4 Covariates association with the timing of interventions

The treatment here is the timing of either stay-home orders or 50% mobility reductions, where the time is expressed in epidemic time (days since 3 deaths per 10M people). We examined the residuals of a linear regression to assess the associations between the timing of stay-home orders or 50% mobility reductions measured in epidemic time with the NCHS indicators, and the percent of residents that are Black, Hispanic, aged 65 years and older, and attending college. An R-squared of 0.45 (0.3) is obtained when regressing the epidemic timing of stay-home orders (50% mobility reductions) with state random effects. All covariates are significant except for the proportion of residents attending college. We validated that the variability and magnitudes of the residuals do not change much as a function of the fitted values, indicating there is a reasonable linear relation between the treatments and covariates.

Geographic dependence is presumed in the epidemic timing of stay-home orders which have a Moran’s I spatial auto-correlation of 0.35 and of 0.40 in the case of the 50% mobility decrease. To learn whether the covariates capture the spatial dependence of the treatments, we obtained the Moran’s I correlation of the linear regression residuals of the timing of stay-home orders and 50% mobility reductions, which have an auto-correlation of 0.003 and 0.007 respectively. This means covariates explain most of the spatial process of the treatments.

**Table S3:** The spatial auto-correlation of the treatment, the epidemic timing of stay-home orders and 50% mobility reductions, is captured by the county-specific demographics and state random effects

|  | **Spatial auto-correlation**<br>**(sd)** | **Spatial auto-correlation**<br>**of residuals (sd)** |
| --- | --- | --- |
| Timing of stay-home<br>orders epidemic timing | 0.37 (0.03) | 0.03 (0.03) |
| Timing of 50% mobility<br>decrease epidemic timing | 0.23 (0.03) | 0.007 (0.03) |

### B Details of the Statistical Modeling Approach

#### B.1 A Two-stage Model for the Post-Intervention Trend Shift in Death Trajectories

There are two important features that go into the definition of a death trajectory. First, for each county, time is measured in *epidemic time*, which, in contrast to *calendar time*, is defined as the number of days elapsed since local arrival of the epidemic. For this study, such date is considered as the day in which the county reached a deaths threshold of 3 deaths per 10 million residents. Second, because reports of daily deaths often exhibit erratic day-to-day variation across counties (e.g., one county may report most cases on Monday while another county on Wednesday), the analysis uses 7-day centered rolling average of daily deaths, which allow to remove day-of-week reporting effects.

Let *y*_*it*_ be the moving average of daily deaths observed in county *i* at time *t*, and let *N*_*i*_ be the county’s population. At the top layer of the hierarchical model, the distribution of *y*_*it*_ is described using a negative binomial regression, which is a standard choice for count data with high variance

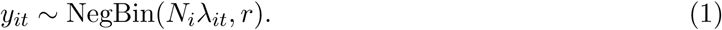

With this parameterization, 𝔼 [*y*_*it*_] = *N*_*i*_*λ*_*it*_ and 𝕍[*y*_*it*_] = *N*_*i*_*λ*_*it*_(1 + *N*_*i*_*λ*_*it*_*/r*). The unknown parameter *r* indicates over-dispersion: the smaller it is, the higher the variance. And as *r* → ∞, the distribution of *y*_*it*_ converges to a Poisson distribution, which has the least possible over-dispersion. The population *N*_*i*_ is used so that *λ*_*it*_ can be interpreted as a *per-capita* rate.

The next layer of the model, comes from the specification of the *per-capita* rate *λ*_*it*_, defined in terms of a pre- and post-intervention term. Each one of these terms are functions of time, covariates, and random effects that capture the spatio-temporal nature of the process as well as county-specific heterogeneity. Let ***x***_*i*_ be the set of county-level features (i.e, NCHS category and demographics) and *timing* _*i*_ be the timing of the intervention at county *i* (i.e., the days between reaching the threshold deaths and the intervention). Then, the per capita rate is modeled as

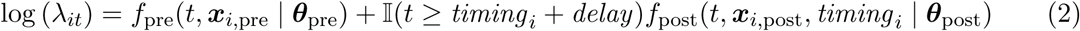

where the quadratic trend functions (that will be defined in detail below) *f*_pre_ and *f*_post_ depend on the unknown parameters ***θ***_pre_ and ***θ***_post_, as well as a set of covariates for each county ***x***_*i*,pre_ and ***x***_*i*,post_. An important remark about equation 2 is that the post-term (the term involving *f*_post_) is zero for all times before the intervention (plus delay), whereas the pre-intervention term is defined for all *t*’s before and after the intervention; equation 2 works similar to a spline, except that the terms are additively composed. To obtain some intuition, one may wish to think about the term with *f*_pre_ as the trend under no intervention whatsoever, and the post-term involving *f*_post_ as the *change* in trend due to the intervention. However, we raise some caution against such a strong causal interpretation, which requires additional considerations. Figure 2 shows a visual schematic of the model for further intuition. Note that equation 2 is well defined even in the absence of an intervention (in which case the second term vanishes).

#### B.2 Covariates interactions with time polynomials

For ***x***_*i*,pre_—the covariates for the pre-intervention trend for each county in equation 2—the variables included are dummies for the NCHS category (with the NCHS 1 as the control group) and county-level demographics (percentage of black, white and hispanic population, as well as percentage with college degree). To maximize the expressivity of the model, we also included the interaction terms between the NCHS category and the county-level demographics.

For ***x***_*i*,post_—the covariates for the post-intervention trend for each county in equation 2—we also include the NCHS dummy variables and the county-level demographics, but we did not include interaction terms between these to avoid over-fitting. As noted in equation 3, the post-intervention trend also includes a variable for the days between the intervention and the deaths threshold, and this variable also interacts with the linear and quadratic term of the post-intervention time polynomial.

To improve stability when fitting the model and make weakly informative priors comparable, all covariates are standardized by their mean and standard deviation.

#### B.3 Specification of the Pre- and Post-intervention Trend Functions

We now specify the functional forms of *f*_pre_ and *f*_post_. First, for *f*_pre_ we use a quadratic polynomial where the coefficients are obtained from county-level covariates and random effects. First, we introduce necessary notation. Let (1, *t, t*^2^)^*T*^ denote a 3-variate vector with a factor of the epidemic time *t* for each polynomial degree. The pre-trend factor contains both fixed (regression) effects and random effects for each polynomial degree. For the regression coefficients, first we consider a 3-variate intercept ***α***_pre_ with a value for each polynomial degree, and a matrix of coefficients ***β***_pre_ with one column for each polynomial degree and one row for each value of a subset ***x***_*i,pre*_ of the county features. Now let ***u***_*i*_ be the 3-variate random effect for county *i* that captures county-level heterogeneity and spatial effects. And finally, let 𝒱_*it*_ denote a *temporal* effect that captures autocorrelated variation around the main trend. This last-term does not depend on the polynomial degree, but instead there is one for each observation *y*_*it*_. Using the newly introduced variables ***θ***_pre_ = (***α, β***_pre_, ***u***, *𝒱*), the pre-intervention trend function is defined as

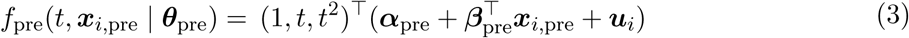

The random effects ***u***_*i*_ are composed of three elements:

1. state-level effects to capture correlation due to state-level policies and outcomes;
2. a spatial component for the remainder spatial correlations not captured by state-level effects;
3. a heterogeneous component for county-level unstructured variation.

Further details about the random effects are given in the next section. We now explain the design of the post-intervention trend, which is also defined as a quadratic polynomial term with regression coefficients, but only containing fixed effects ***θ***_post_ = (***α***_post_, ***β***_post_). One major difference is that the polynomial term is shifted so that is is exactly zero at the day of the intervention plus the lag term capturing the delay from infection to death

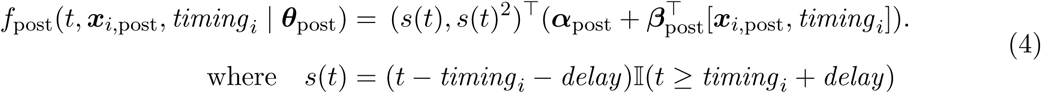

We remark two things about the post-trend. First, the regression coefficients for the post-trend explicitly contain an additional term corresponding to the timing of the intervention to allow for differentiated slopes for early and late intervention. Second, note the omission of the zero-degree polynomial term. This is done to guarantee the continuity of *λ*_*it*_ before and after the intervention. The regression coefficients ***α***_pre_, ***α***_post_, ***β***_pre_ and ***β***_post_ are given weak priors *N* (0, 15). The covariates ***x***_*i*_ are standardized to avoid scaling issues.

#### B.4 Specification of the State, Spatial and Unstructured Random Effects

The county-specific random effects ***u***_*i*_ for county *i* are composed of three terms

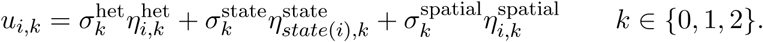

where *k* stands for the polynomial degree and we use state(*i*) to indicate the state that county *I* belongs to.

To specify priors, we adapt the popular Besag-York-Molli é model^18,20^. Let 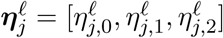 where, *𝓁* ∈ {state, county, spatial}. Then the prior for the state and county-level heterogeneous component are given by

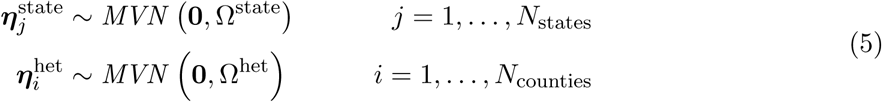

where Ω^state^ and Ω^het^ are random correlation matrices. The correlation matrices Ω are given LKJ(1.1) priors^21^ which are weakly informative. For the spatial effects, we use an intrinsic conditional autoregressive term (ICAR) where each term is conditionally dependent on weighted values of its neighbors

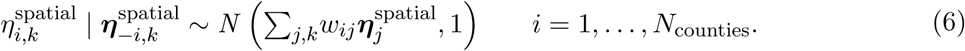

with the weights *w*_*ij*_ indicating a weighted average from the neighbors of the county determined by the adjacency matrix. The weights are scaled proportional to the average number of neighbors in each connected component in the graph, similar to (^18^), to approximately encourage that the marginal variances in all connected components are comparable. For constructing this adjacency matrix, a flexible specification is necessary since most counties in the dataset have no spatial neighbors (roughly 75%, see figure 1). Thus, we merge the edges from the geographical adjacency matrix with the 2-nearest-neighbors graph with a maximum radius of 100kms. Including the nearest neighbors greatly reduces the disconnectedness of the graph.

Since we regard the state-level effects as actual controls in the regression model, we give their scale a weakly informative prior 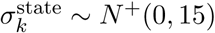 that is comparable to the priors of the coefficients of other covariates in the model. For the scales of county-level effects, it is important to choose a fair prior balancing the variances of the heterogeneous and spatial components. Thus, we use the following priors

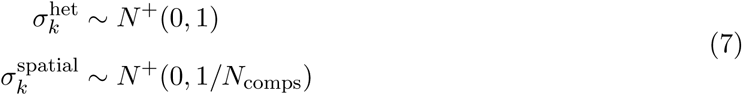

where *N*_comps_ the number of connected components in the graph determined by the adjacency matrix.

#### B.5 Counterfactuals

We base inference on simulations from the posterior distributions of the parameters in models of the form (4). In the simulations the intervention timing, *d*_*i*_, is replaced with alternative hypothetical timing corresponding to earlier or later intervention. Specifically, under different values of *d*_*i*_, we take a sample from the posterior predictive distribution for each county and take group averages by NCHS category for each time *t*. Group-average curves are calculated by averaging point-wise posterior predictive quantities among counties within the same NCHS category. Differences in cumulative deaths across counties shown in 1 were calculated by aggregating posterior-predicted daily deaths for all counties in an NCHS county category, on a given date and intervention-timing scenario, then accumulating resulting values across days through April 20 and comparing the resulting predictions under the observed vs. early intervention scenario and the observed vs. late intervention scenario.

#### B.6 Modeling the Unknown Distribution of the Time from Infection to Death

In this variant of our statistical model, we seek to evaluate both the sensitivity to fixing the delay from infection to deaths at 14 days. For this purpose, we treat the delay as another parameter of the Bayesian hierarchical model and put a prior to it to represent existing research about it.

First, we remind a few facts about scientific knowledge about the delay. Lauer et al.^15^ estimate the first quartile of time between infection to symptoms to be 3.8 days, while Yang et al.^16^ estimate the first quartile of days between symptoms to death at 10 days. By adding both we obtain an approximation of the left tail of the distribution of time lag of deaths at 13.8 days. A similar calculation is done by Wilson et al.^17^ using IQR values. Using an analogous reasoning for the medians, the median time between infection to symptoms is close to 5.1 days^16^ and the median of days between symptoms to death is around 12 days^15^. Therefore, 17 days is close to the median time from infection to death, time at which the trend shift should be highly visible.

We use this information to specify a reasonable Bayesian prior on the delay. We propose to use a scaled and translated Beta(2, 2) prior in the range [10, 18] with parameters (2, 2), so that the mean is 14 days. This prior is shown in Figure S10 in the detailed results section C.7 of this Web Appendix together with the posterior distribution after fitting the model.

We remark that the purpose of this model is not to substitute the model presented in the previous sections, but rather to assess the sensitivity to the specification of the 14-day delay between an intervention and the trend shift in the main model. It will be shown that the model conclusions are not highly sensitive to the choice of 14-days.

### C Results and Model Validation

#### C.1 MCMC convergence statistics

Figure S4 shows the Gelman-Rubin 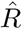 convergence statistic for the fitted stay-home model^22^. The figure shows that all main effects are within ideal (heuristic) values (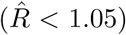) indicating convergence of the chains. In the case of the random effects (heterogeneous and spatial), only a very tiny fraction exceed the recommended threshold (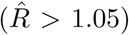). Inspecting a few of these cases, we found that they correspond to small counties with few (mostly zero) observations, and mainly to the coefficient for the quadratic term of the trend polynomial. Overall, the figure suggests good convergence of the chains. Figure S5 complements this analysis with traceplots for the ***β***_post_ coefficients. Other main coefficients have similar behavior. The behavior for both figures is also extremely similar for the mobility decrease model.

**Figure S4:**
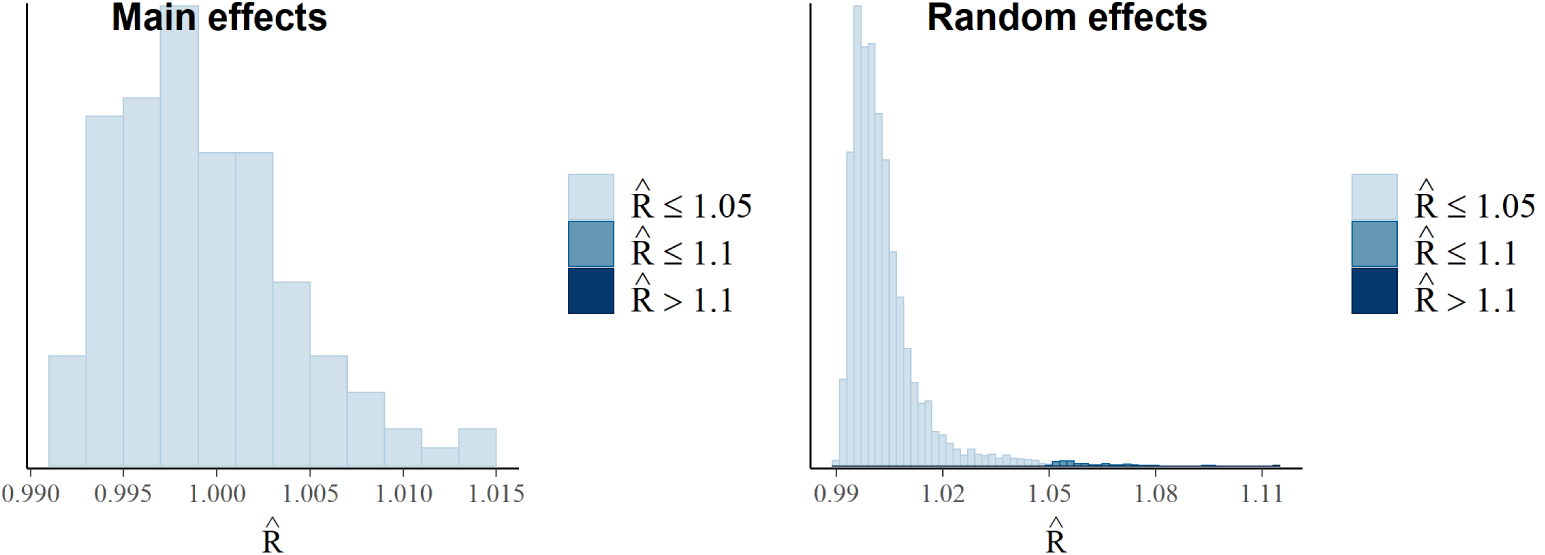
Gelman-Rubin convergence statistic.

**Figure S5:**
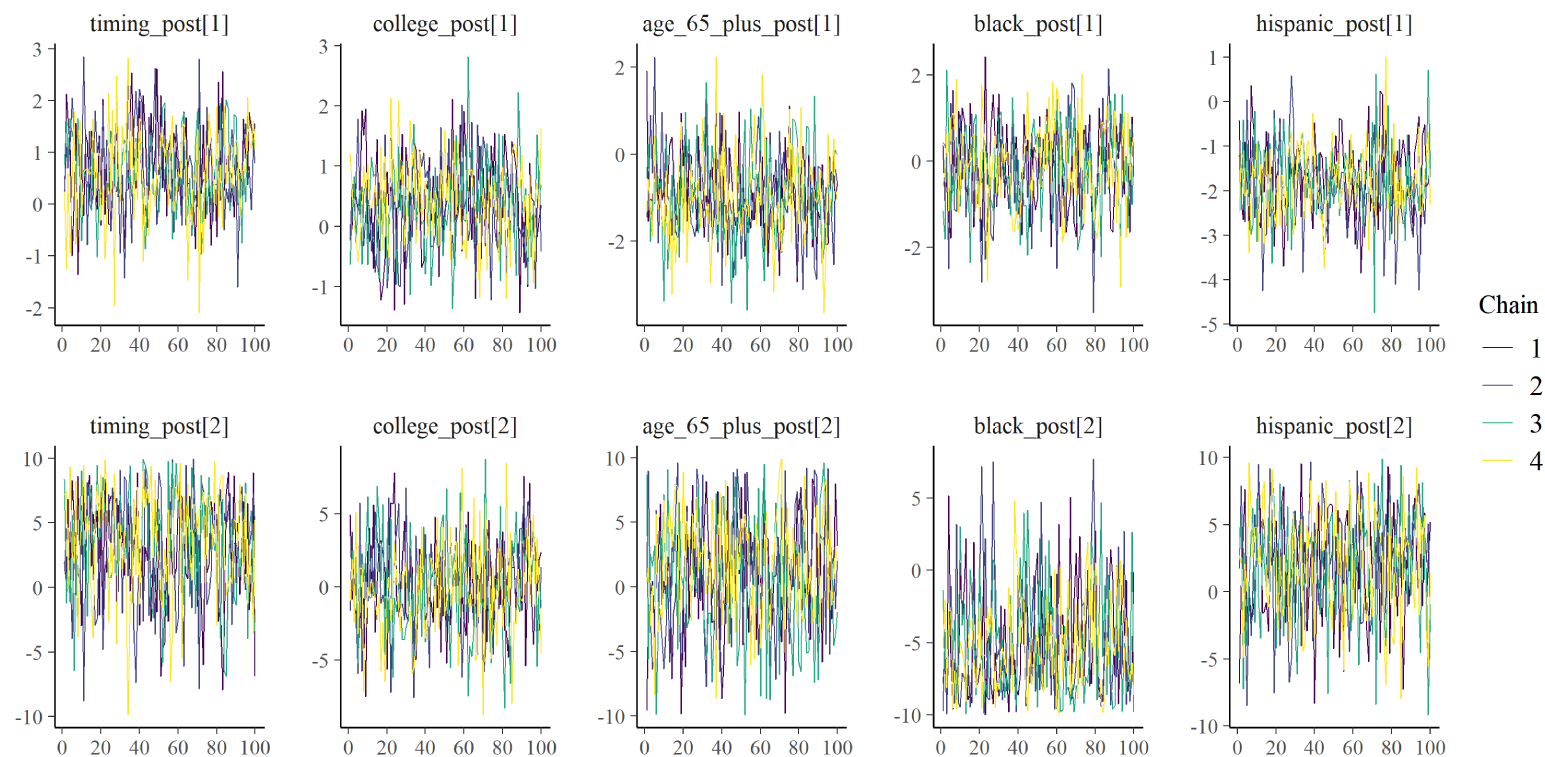
Traceplot for ***β***_post_ showing similar distribution for all chains. Samples are after warmup for 1000 chain iterations with a thinning of 10.

#### C.2 Goodness-of-fit

We conducted tests to evaluate the fit of the models. Overall, the tests in this section allow to conclude that the proposed statistical model with a two-stage quadratic trend does a good job a fitting the data. This will be further illustrated in section C.6, that shows examples of fitted curves for counties with a large number of observed deaths.

First, we note that a fraction of approximately 82% of the posterior draws are the equal to the observed value (in both the stay-home and decrease model). This is expected since most observations are zero for small counties. For the data corresponding to the top 25 counties with the most deaths, the fraction of posterior draws that are exactly equal to the observed data are 28% and 26% for the mobility and stay-home models respectively. These quantities can be interpreted as the coverage of zero-width confidence intervals. We also evaluated the coverage of the posterior quantile-confidence intervals for these top counties. The 10%, 50% and 90% (centered) quantile-intervals for the mobility model cover 45.5%, 87% and 99% of the data respectively. For the stay-home model, the same quantile-intervals cover 45.6%, 88.6% and 99.2%.

These coverage of the confidence intervals appears to be overly conservative. In general, it is preferable to be conservative about the estimation of correlations between the timing of interventions and death trajectories. However, the intervals are a bit difficult to interpret given the high precision of the zero-width intervals. To offer more intuition, we also evaluate the following metric for relative error

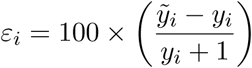

where 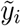 is a random sample- from the posterior predictive distribution. The metric is the prediction error or residual in relative percentage terms—up to adding one in the denominator to ensure it is defined when *y*_*i*_ = 0, and which is negligible for large values of *y*_*i*_. The results are shown in Table S4. The table demonstrates that the median relative error is 0 for both models (mobility and stay-home), even when considering only the 25 counties with the most deaths. This shows that the model is unbiased (in the log-scale). The table also shows that 50% of all posterior draws (measured by the IQR) are within 20% of the observed data, which is surprising given the rigid form of the quadratic trends—indicating that the two-stage approximation captures the data well. The quantity is only computed for the 25 counties with the most deaths since the relative error is even lower when considering all counties, since it is mostly 0% due to the large number of zeros as previously remarked.

**Table S4:**
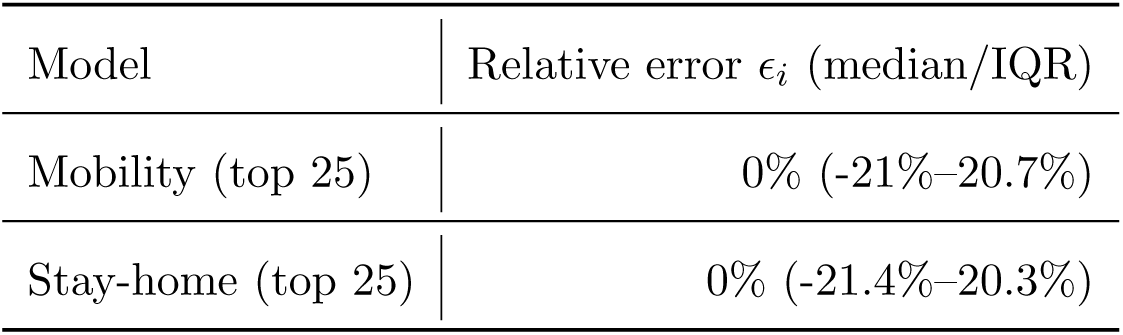
Relative error of the posterior predictive distribution. The error is computed for each observation and each posterior draw. The right column shows the median of all draws and observations with the IQR (25% and 75% quantiles) in parentheses.

#### C.3 Spatial Analysis of Model Residuals

The goal of this/ section is to perform a sanity check of the model residuals. We verify that the residuals are not spatially correlated. For this analysis we condition on the time since the deaths threshold to further make sure the residuals do interact with the temporal component. We use Moran’s I as the measure of spatial correlation, computed from the inverse-distance-weighted adjacency matrix between counties. The residuals considered for the analysis are the median deviance residuals. Figure S6 shows the results from Moran’s I for the stay-home model for each time, suggesting no evidence of spatial correlation. The results for the mobility-model are very similar.

**Figure S6:**
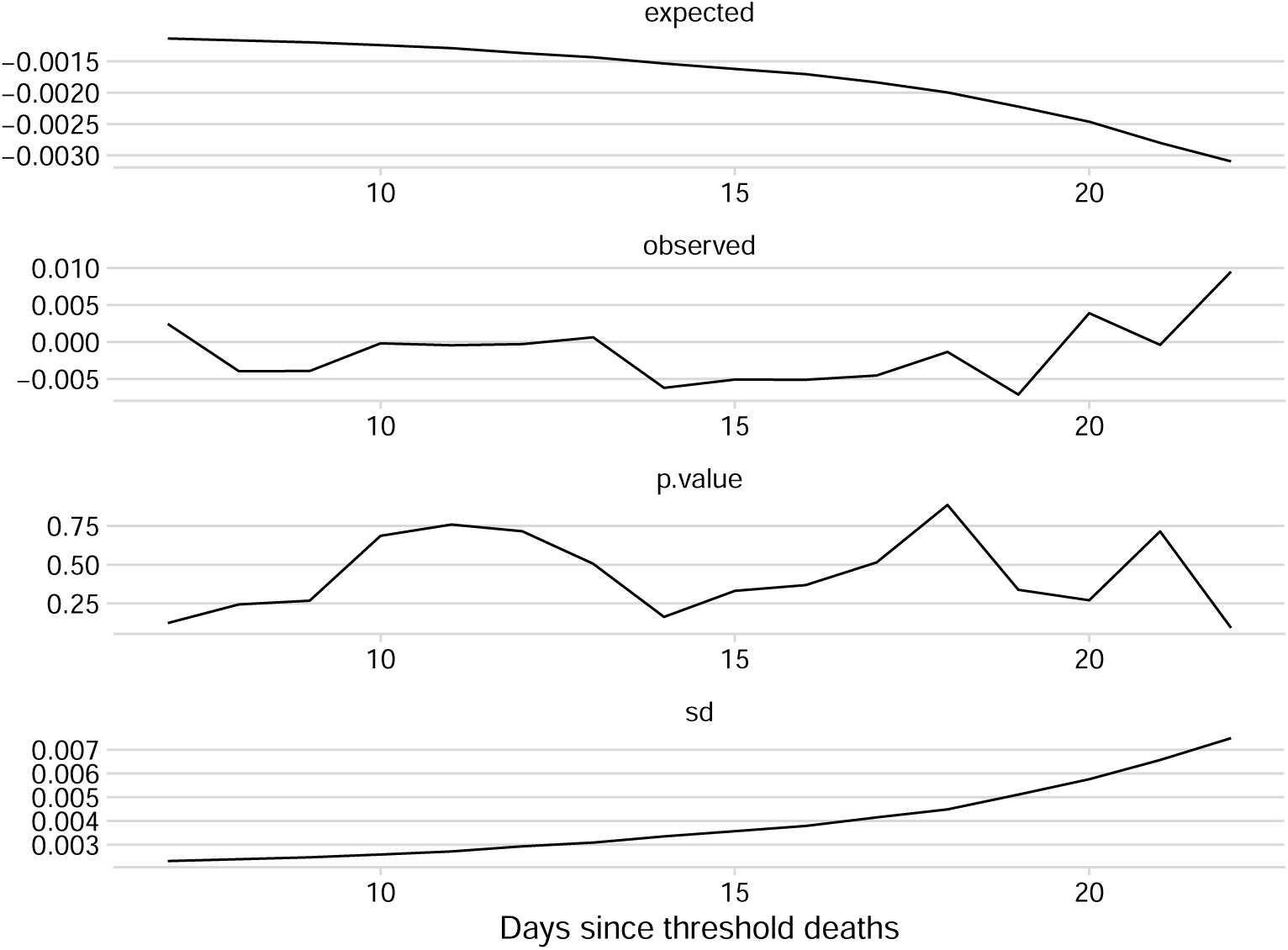
Moran’s I spatial correlation analysis for the stay-home model’s deviance residuals for different times since the deaths thresholds.

#### C.4 Robustness to the Exclusion of New York City Counties

We conducted an analysis to validate the model fit sensitivity with respect the counties that concentrate the largest number of deaths. For this test, we remove the data from the seven counties within the New York City area, all which are within the top 10 counties that concentrate that largest number of deaths in the period of study. These counties are: Queens, New York, Kings, Nassau, Richmond, Suffolk and Bronx. After removing these counties from the data set, we compared the predicted excess/averted deaths for both models. Figure S7 shows this comparison for the stay-home model. Looking at the figure, it is readily apparent that removing those counties did not change the fit and estimates of counterfactuals for the remaining counties. The equivalent figure for the decrease model is very similar. Our judgment is that the negative binomial accommodation of over-dispersion and the implicit regularization through the random effects provides sufficient protection against results being dominated by a small number of large counties.

**Figure S7:**
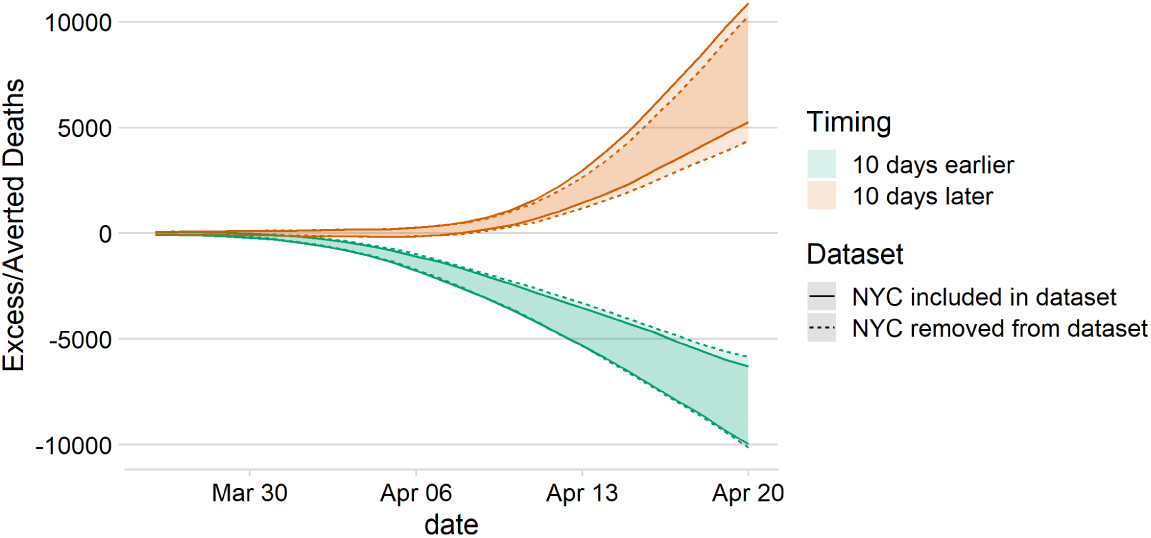
Comparison of the predicted excess/averted deaths in all but the 7 counties from NYC when removing those 7 counties from the data set. In the solid line, the counties are used for fitting the model but removed from the computation of counterfactuals. In the dashed lines, the counties are not included while fitting the model. The comparison shows the sensitivity of the model to those data points. Results are for the stay-home model.

#### C.5 Out-of-sample Post-Intervention Prediction for Large Cities

We performed a short experiment to assess the quality of the model for predicting a post-intervention bent. For this task, we removed from the data set all the post-intervention data (accounting for the delay from in infection to death) from four counties among the top 10 counties that concentrate most of the observed deaths: Queens (New York City), Los Angeles (California) and Wayne (Michigan). Only pre-intervention data from these counties are included, which is used during model fitting to obtain an approximation of their county-specific random effects. However, their post-intervention data in this experiment are unobserved. We then evaluate the predictive power of the model using the held-out post-intervention data for these counties as validation. The results are shown in Figure S8. Quite strikingly, most predicted observation fall within the predicted 95% confidence intervals, which shows that the predictive power of the model is quite good. In particular, the median prediction is very accurate for Queens County in New York, which can be hypothesized is due to the high degree of spatial correlation between neighboring counties whose post-intervention data was included for fitting the model in this experiment..

**Figure S8:**
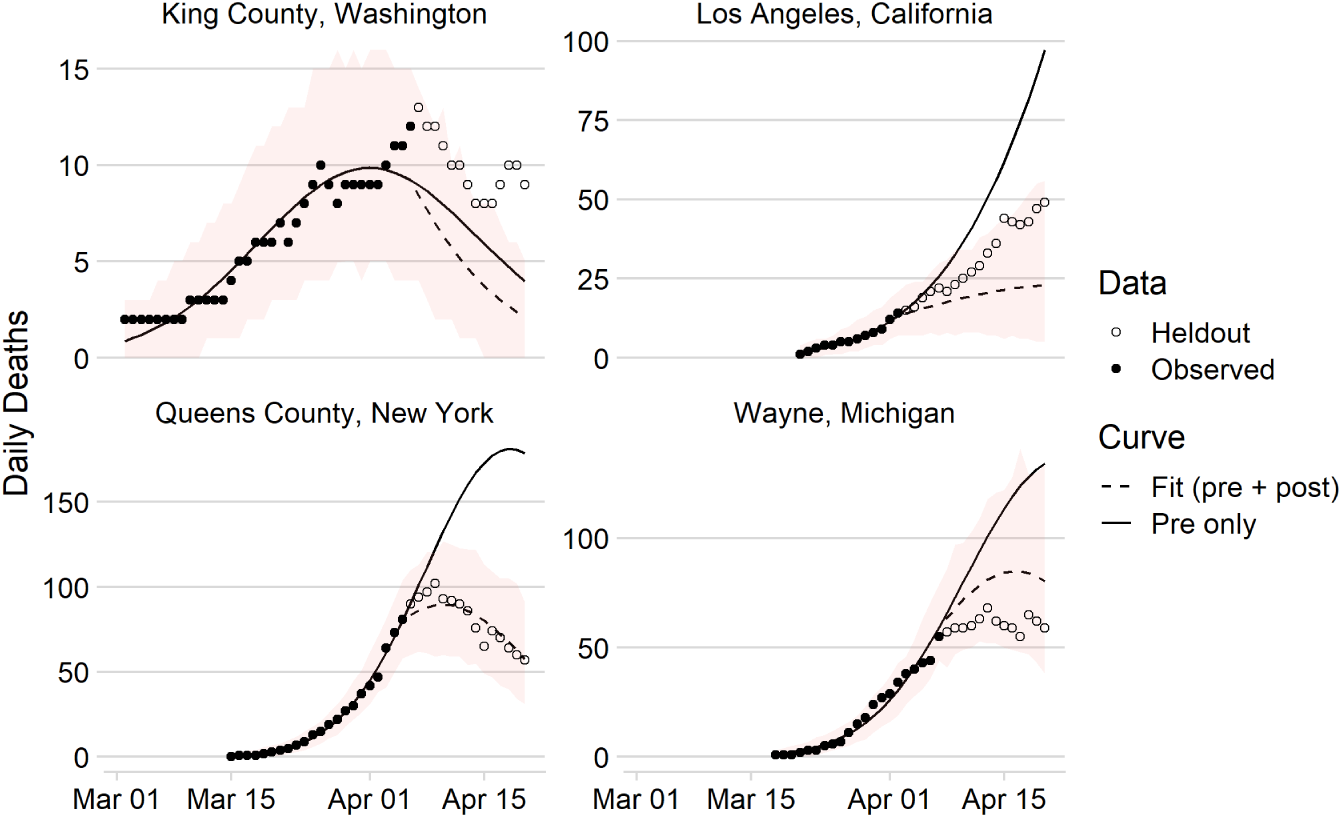
Validation of the stay-home model’s predictive power for heldout post-intervention data from four counties with many observed deaths in the period of study.

#### C.6 Examples of County-level Fitted Curves

Figure S9 shows the observed and hypothetical trajectories for six example counties: King County, Washington; Queens and New York Counties, New York; Jefferson County, Louisiana; Wayne, Michigan; and Los Angeles California. In King County, Washington, a hypothetical late adoption of a stay-home order does not appear to have a significant impact on the post-intervention death curve trajectory, suggesting that its curve had already flattened by the time the policy was adopted, likely as a result of awareness brought by national attention as the first US cases emerged in this county.

#### C.7 Results of the Random Lag Model

Figure S10 shows the results of the model that estimates the delay from infections to death. The figure shows the prior and posterior distribution. The figure shows that for both the Stay-home and Mobility decrease models the posterior mode approximately concentrates in the range 14-15.

**Figure S9:**
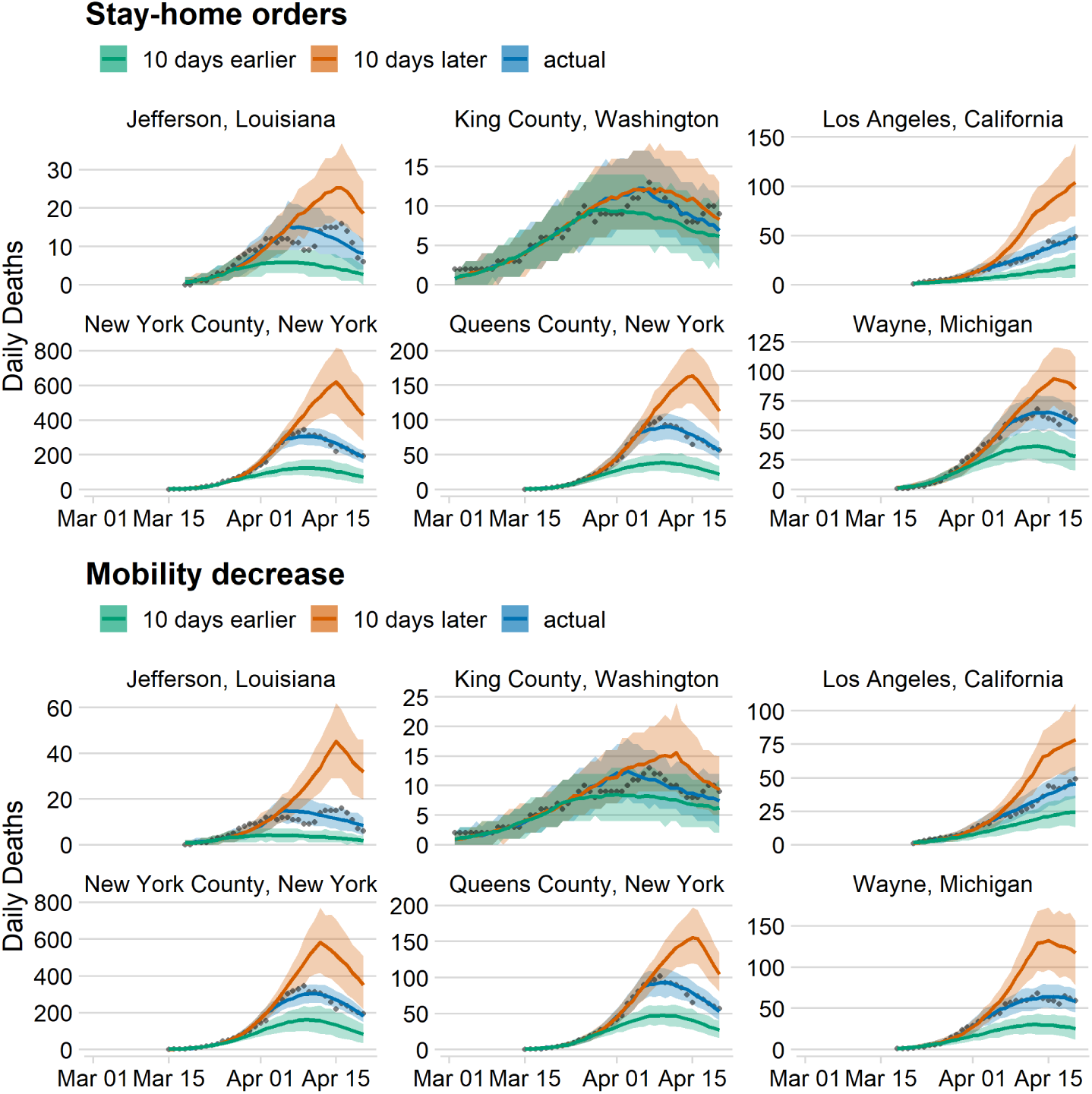
The early and late intervention timing counterfactuals for six example counties. The median and 90% credible intervals are shown for each county and each intervention type.

**Figure S10:**
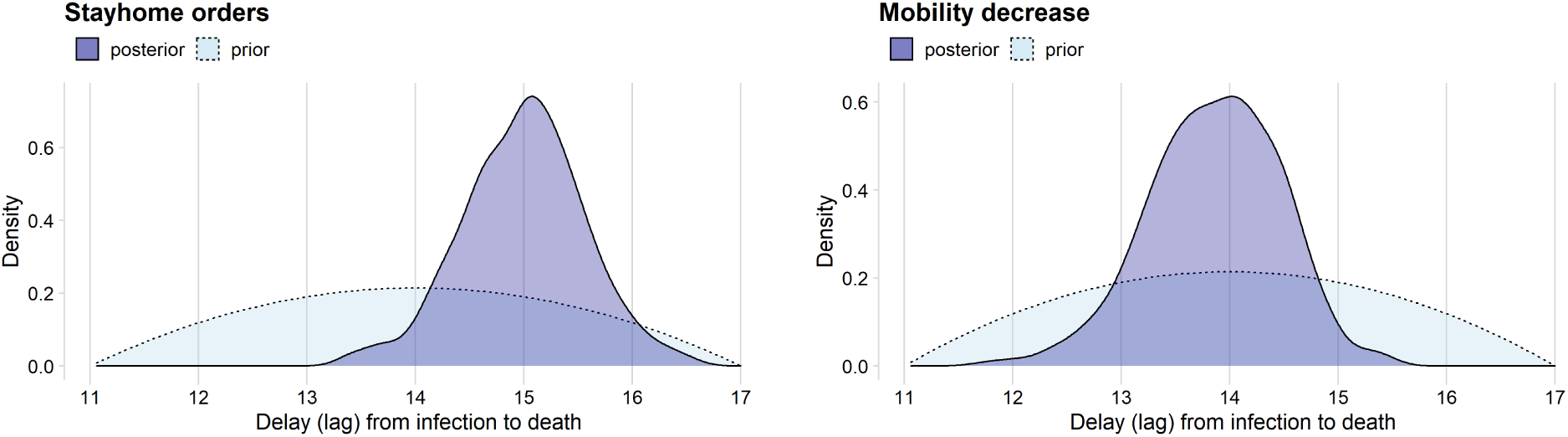
Prior and posterior distribution from the random changepoint model.

